# DEFINITION AND CLINICAL PRACTICE OF MANUAL THERAPY IN TRAUMATIC SPINAL CORD INJURY: A SCOPING REVIEW PROTOCOL

**DOI:** 10.1101/2025.05.06.25327077

**Authors:** Maini Irene, Bonavita Jacopo, Casonato Caterina, Feller Daniel, Trentini Francesca, Zarbo Michele, Zadra Alessandro

## Abstract

**Introduction:** Manual therapy has been a topic of growing interest and debate within the musculoskeletal field, particularly concerning its definition, mechanisms, and implementation in clinical trials. It has also emerged as a rehabilitative intervention for spasticity, often used in combination with pharmacological treatments, particularly in stroke populations. However, individuals with spinal cord injury (SCI), who commonly experience spasticity, have received less attention in this context, despite the significant impact of spasticity on their independence and quality of life.

**Objective:** This scoping review aims to explore how manual therapy is defined and implemented in the treatment of spasticity among adults with traumatic SCI. Specifically, it seeks to identify which interventions are labelled “manual therapy”, how spasticity and outcomes are defined, and whether patient-centered goal setting is incorporated. It also evaluates the quality of intervention reporting using the TIDieR checklist and investigates the feasibility of conducting a future systematic review.

**Methods:** The review will follow JBI methodology and be reported according to the PRISMA-ScR checklist. Studies in English including adults with traumatic SCI and spasticity will be selected from MEDLINE, EMBASE, and Web of Science. Independent reviewers will perform screening and data extraction, with a descriptive synthesis of the findings.

## INTRODUCTION

Manual therapy has recently become the subject of literature in the musculoskeletal field, with debates arising regarding its definition and conceptual paradigm [1], the mechanisms underlying its therapeutic effects [2], and the actual nature of interventions described as “manual therapy” in clinical trials [3].

Manual therapy has emerged among the interventions reviewed for treating spasticity in individuals post-stroke, often used in synergy with intramuscular focal treatments [4]. Some studies have explored its influence on instrumental measures of spasticity in this population [5].

Various rehabilitative interventions, including manual therapy, have been recently reviewed in individuals with spastic equinovarus foot following stroke [6].

People with spinal cord injury (SCI), similarly to stroke survivors, often present with a high prevalence of spasticity. The literature widely documents the impact of spasticity on independence and quality of life [7,8,9]; therefore, we chose to focus our attention on this specific population.

## OBJECTIVE

We developed our research question using the Population-Concept-Context (PCC) framework as follows:

- Population (P): Adults (age > or = 18 years old) with traumatic SCI affected by spasticity, with no limitations in gender, level, or completeness/incompleteness of the lesion.
- Concept (C): Manual therapy interventions, applied by any healthcare professional, even in combination with other treatments.
- Context (C): Any.

We thus formulated the following research question:

“What is the definition of manual therapy adopted in the treatment of individuals with traumatic SCI affected by spasticity? Which rehabilitative interventions described in the literature are labelled as ‘manual therapy’ in this population?”

Additionally, we wanted to address how spasticity is defined and measured, the quality of intervention reporting, and how treatment goals are established and negotiated within this context.

Given the exploratory nature of this question, a scoping review design was chosen, aiming to explore the current literature on this topic and provide a critical, descriptive (not quantitative) synthesis.

### Primary objectives

For each included study:

- Report the definition of “manual therapy” adopted.
- Identify which rehabilitative interventions administered match the definition of “manual therapy”.

### Secondary objectives

For each included study:

- Report the adopted definition of spasticity.
- Report the outcome measures used.
- Assess the quality of intervention reporting using the TIDieR checklist [10].

Regarding goal setting, for each included study we will answer the following questions:

- What treatment goals were defined?
- Were time horizons for those goals specified?
- Who defined the goals?
- Was the person with traumatic SCI involved in goal definition?
- What is the reported efficacy of the intervention?

We will also determine whether enough RCTs were found, homogeneous in population and intervention, to support a future systematic review.

## MATERIALS AND METHODS

This scoping review will be conducted in accordance with the “Joanna Briggs Institute” (JBI) methodology for scoping reviews [11]. The PRISMA-ScR checklist (Preferred Reporting Items for Systematic Reviews and Meta-Analyses extension for Scoping Reviews) will be used for reporting [12].

### Inclusion criteria

- Primary studies evaluating a rehabilitative intervention delivered by any healthcare professional.
- Studies in which most of the population (>50% of participants) consists of adults with traumatic spinal cord injury and spasticity, excluding those with traumatic brain injury.

### Exclusion criteria

- Studies in which the full text does not include the specific phrase “manual therapy” to describe at least one of the considered rehabilitative interventions.

### Sources

All primary studies in English will be included, with no limitations regarding date, geographical area, or setting. Abstracts, posters, and studies without full-text availability in English will be excluded.

Duplicate removal will be performed using the dedicated “Systematic Review Accelerator” tool [13].

### Search strategy

Searches will be performed in the following databases: MEDLINE (via PubMed), EMBASE, and Web of Science.

The MEDLINE search will use the following string:

*(“Muscle Hypertonia”[MeSH Terms] OR “Muscle Tonus”[MeSH Terms] OR “Spasm”[MeSH Terms] OR “Contracture”[MeSH Terms] OR “spasm*”[Title/Abstract] OR “spastic*”[Title/Abstract] OR “hyperton*”[Title/Abstract] OR “hypermyoton*”[Title/Abstract] OR “hyperreflexia*”[Title/Abstract] OR “reflex, abnormal”[MeSH Terms] OR “Muscle Tone Increased”[Title/Abstract: ~2] OR “increased muscle tone”[Title/Abstract: ~2]) AND ((“Spinal Cord Injuries”[MeSH Terms] OR ((“spine”[Title/Abstract] OR “medulla*”[Title/Abstract]) AND (“injur*”[Title/Abstract] OR “traum*”[Title/Abstract] OR “contusion*”[Title/Abstract] OR “laceration*”[Title/Abstract] OR “lesion*”[Title/Abstract] OR “compression*”[Title/Abstract] OR “fracture*”[Title/Abstract]))) NOT (“Animals”[MeSH Terms] NOT “Humans”[MeSH Terms])) AND (“manual therap*”[Title/Abstract] OR “mobiliz*”[Title/Abstract] OR “mobilis*”[Title/Abstract] OR “manip*”[Title/Abstract] OR “Musculoskeletal Manipulations”[MeSH Terms] OR “Physical Therapy Modalities”[MeSH Terms] OR “manipulation, chiropractic”[MeSH Terms] OR “rehabilitation”[Title/Abstract] OR “physiotherapy”[Title/Abstract] OR “physical therapy”[Title/Abstract] OR “stretching”[Title/Abstract] OR “Muscle Stretching Exercises”[MeSH Terms] OR “occupational therapy”[Title/Abstract])*

### Study selection

Two reviewers will independently screen studies first by title/abstract and then by full text. Discrepancies will be resolved by consensus or third-party adjudication. The Rayyan QCRI platform will be used to organize and monitor the selection process.

### Reference searching

Reference searching will include both backward and forward citation tracking for each included study, in order to identify additional relevant records cited by or citing the included works.

### Data extraction

Data will be extracted using two templates developed in Microsoft Word. The extraction form will be pilot tested in three studies. Data extraction will be conducted independently by two reviewers. Any discrepancies will be resolved by consensus or, if needed, by a third reviewer.

### Data synthesis

Findings from the scoping review will be summarized descriptively using tables and diagrams. Extracted data will be collated and synthesized to assess whether enough homogeneous RCTs are available to support a future systematic review.

### Availability of data

All data generated from this work will be available in the final manuscript.

## Data Availability

All data produced in the present study are available upon reasonable request to the authors

## Notes

### Competing Interest Statement

The authors have declared no competing interest.

### Funding Statement

This study did not receive any funding

